# The use of a new type of heart rate-controlled training system “HeartGo ®” for patients with chronic Heart failure on the pedelec

**DOI:** 10.1101/2020.08.17.20157867

**Authors:** Erik B. Friedrich, Herbert Löllgen, Helmut Röder, Wolfgang Baltes, Oliver Adam, Martin Schlickel, Günter Hennersdorf

## Abstract

**Background:** The study “HI-Herz.BIKE Saar” (August 2017 - September 2019) examined the health benefits and training effect of e-bikes (pedelecs) in patients from ambulatory heart groups with moderate chronic heart failure (CHI).

**Method:** 10 subjects with NYHA stage II-III and a left ventricular ejection fraction LVEF of <=50% were selected. The presented study is explicitly marked as a pilot study.

The new HeartGo® system used here for the first time allows heart rate controlled training on a special pedelec via a smartphone app. The groups were accompanied during the training rides by a doctor and a paramedic. No cardiac complications occurred. The training units were increased in duration, distance and target frequency every six months.

Frequency behaviour, pedaling and motor load on the pedelec as well as climatic data such as ejection fraction, a biomarker (NT-pro BNP), risk factors, arterial blood pressure and ergometric courses were measured.

**Results:** The power tolerance increased by almost 2.5 times, a discrete decrease of the resting heart rate by 3.7% was observed and pedaling power improved accordingly. In the clinical data the ergometric power increased by 44% and the LVEF improved by 29%. The NT-pro BNP value decreased by 27%. Body Mass Index BMI remained constant at 27 and cholesterol levels showed no significant changes.

**Conclusions:** Pedaling according to this pilot study with its methodological limitations of low number was safe and accompanied by significant health benefits. The subjects were enthusiastic and satisfied with this form of training. This training form can therefore be recommended to heart group participants under certain medical conditions and can be used in the training process. The results of this pilot study with its methodological weaknesses should be verified in a larger follow-up study.

## Introduction

Regular physical activity with endurance and strength training is one of the secondary prevention measures for cardiovascular diseases with high prevalence (1,2). Early rehabilitation should begin in hospital (phase I), be continued in a rehabilitation clinic or as an outpatient close to home (phase II) and then (phase III) preferably in outpatient heart groups (AHG).

A moderate training of 5×30 min or 150 min/week as an effective training goal is aimed at, but often not achieved in heart groups. Endurance sports like walking, running or swimming are recommended. Cycling in heart groups is done less frequently because the performance in the groups is different. Numerous studies have shown that cycling improves endurance, coordination, flexibility and strength; at the same time, the body weight is not carried, so that this sport is also used very well in the case of overweight and is advantageous for heart patients. There is no structured offer for cycling as a rehabilitation sport, especially for patients with heart failure. Here, the recommendations were reserved, since patients with heart failure are usually older participants and because of possible cardiac incidents or accidents, it is assumed that cycling training is dangerous.

Here, electrically assisted cycling is a good option, especially since the pedelec is becoming increasingly popular with seniors. Initial studies show that in overweight people, an improvement in spiroergometric readings is possible using e-bikes (3).

The disadvantage is that older riders usually choose a high pedaling power support because it is “easier” to ride. This reduces the actually desired training effect; there is “gentle riding”.

With a heart rate-controlled system, this relieving posture is avoided. This results in individual stress and comparable loads. This makes controlled endurance training on the pedelec, similar to stationary ergometry, possible as an option in addition to regular training. Studies on this method are not available.

Also a scientific evaluation is then easier to realize, especially since the benefits and risks of regular training of the rehabilitation phase III in outpatient heart groups have not been sufficiently scientifically investigated. Haberecht et al (2013; 4) report on insufficient lifestyle changes and about too little physical activity in heart groups, which is, however, considered sufficient by others (5). It should be noted that participation in a heart group once a week and often less than 45 minutes is insufficient for an efficient training program.

Reports on the use and value of such pedelec assistance systems have not yet been published, apart from some publications on the general use of pedelecs by athletes and seniors with diabetes mellitus (6).

10 subjects with a strictly defined NYHA stage II-III and a LVEF of <=50% were selected. The presented study is explicitly marked as a pilot study, as a new, so far not tested technique was used for exercise control. When selecting the heart failure patients defined in this way, a higher number of subjects could not be reached at only one study centre.

## Methodology

In this pilot study the novel system HeartGo ®, which was tested for practicability by preliminary investigations, was used. This frequency-based system automatically controls the training process. Training and clinical parameters were measured. Training was carried out in summer on level paths along the Saar river (“outdoor” training), in winter inside a sports hall (“indoor” training).

Anthropometric and demographic data at the beginning:

Period 22.8.17 - 10.09.19 with 93 trainings, 31” indoor”, 62 “outdoor”. Number of cases N=10 with a participation frequency of over 90%.

The age was 61.5 (range 43-82) years, 2 participants were female and 8 male, the average BMI was 27 (range 21-37).

The default training or target frequency fraud 96 (range 83-116) beats/min.

For further details see Suppl. 02

## Results

### Technical results

The user-friendliness of the system fluctuated strongly in the beginning. This was fixed within 4 weeks.

At the beginning of the study the stability of the chest sensors also fluctuated. The charge of the used batteries often did not last for the whole training duration; they had to be replaced. The Bluetooth coupling was also not stable. During the duration of the study the disturbances could be minimized.

### Clinical results

The results (blood tests, echocardiography and functional changes) are presented in detail in Tables 3-7 (Suppl. 03).

A questionnaire on well-being before the start and after the end of the study (Suppl.04) showed (Fig. 1A) that subjective well-being improved significantly during the study.

The ratings of percieved exertion (Fig. 1B) (18) on the BORG scale were between 9 and 13, so that subjectively a rather moderate perceived exertion can be assumed. The average rating was 11 = “quite easy”.

**Fig. 1.**
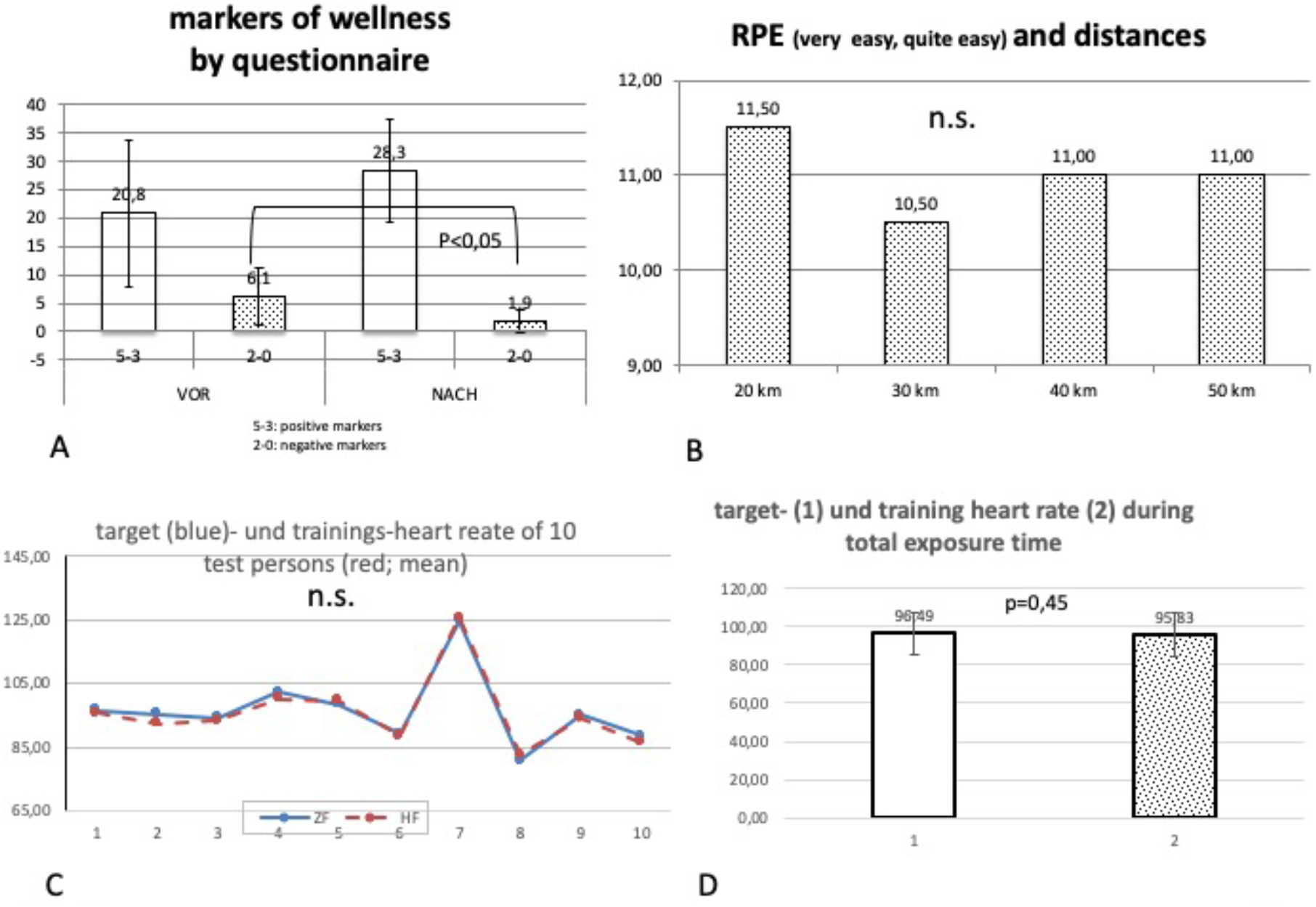

The ratio of target to heart rate of all test persons is shown in Fig. 1 C. There is no difference between the set heart rate (target rate)) and the achieved frequency level of the test persons. Fig. 1 D also shows the nondifferent target and heart rates as overall average values.

The training showed a significant increase in performance:

The ergometry power (in Watt) increased from 91.67 to 132.5 Watt, corresponding to 45%. (Fig. 2A)

The echocardiographically measured LVEF increased significantly from 44.1 to 56.6%. This corresponds to an increase of 29% (Fig. 2B).

**Fig. 2.**
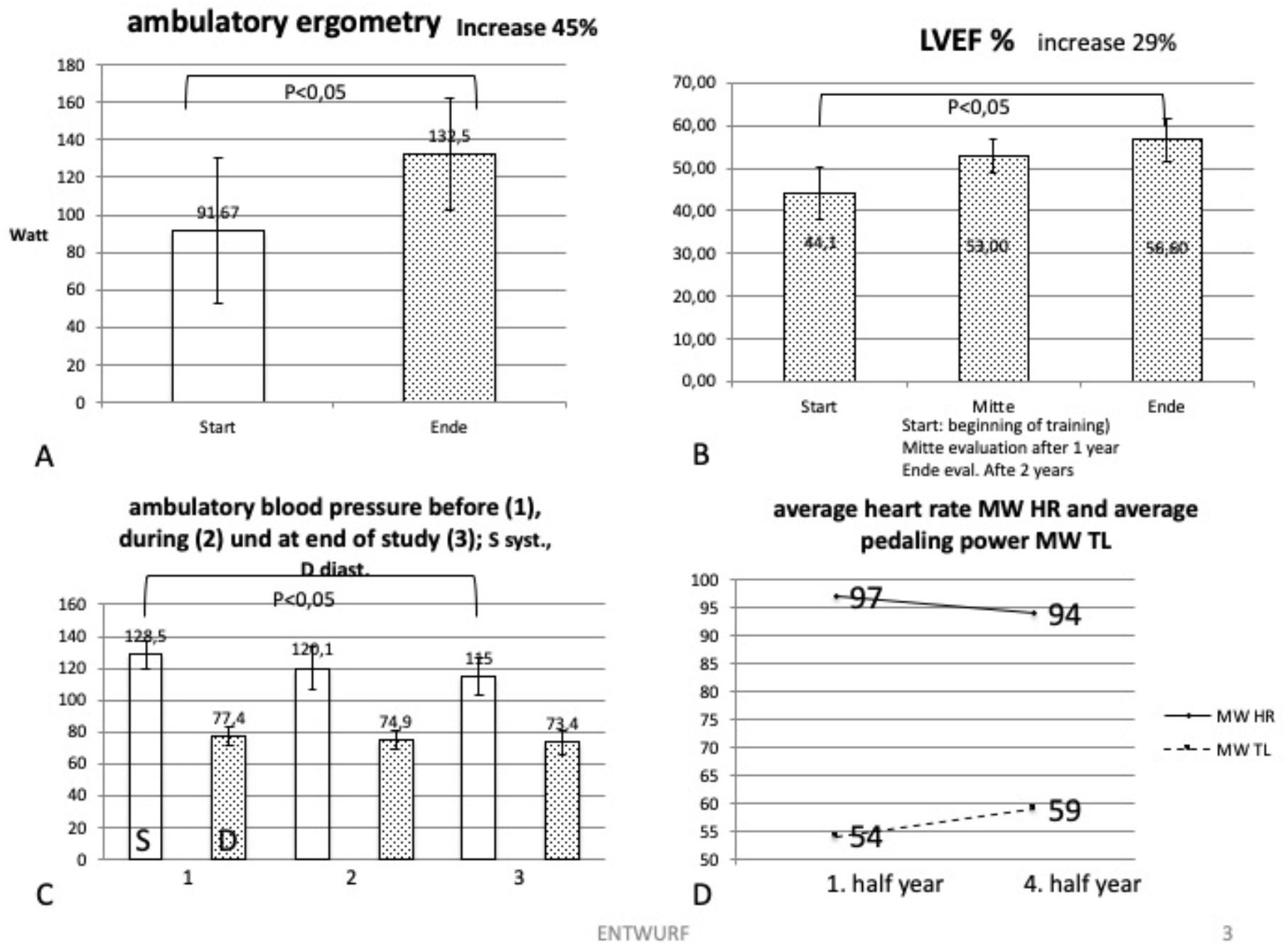

Outpatient systolic blood pressure at the beginning and end of the study decreased significantly from 128.5 to 115 mm Hg (a decrease of 11%) (Fig. 2C)

The pedaling power was 64, the motor power was 79 watts, which is 23% higher than the pedaling power. The latter increased in the 4th half year by about 8.5% compared to the first half year. The resting heart rates showed a decrease of about 4.8% compared to the 1st and 4th half year. Fig. 2D shows this behavior.

### Laboratory parameters

NT-pro BNP: (see Table 3) The initially increased values in all subjects decreased significantly (decrease of 27.2%; p >0.05).

Less noticeable were the values of the lipid status, i.e. total cholesterol, LDL and HDL cholesterol and triglycerides. This only showed trends, but no significant changes. All test persons were undergoing statin therapy at the time of the study.

At the beginning, in the middle and at the end a 6-minute walking test was performed. This showed a non-significant trend towards improvement (increase of 7.3%).

### Complications and side effects during training

At the beginning and at the end of the duration of training there were two incidents (2.15%):

In one patient, a sudden fall from the ride from a bicycle with suspected cardiac syncope. An efficient first aid was given, but also a stay in hospital due to a fracture of the humerus. The cause of the syncope remained unexplained. No consequential damage occurred.

Another subject had an accident with a fall due to an unsafe road (driving over grass). There was also a fracture of the upper arm with hospitalization. No consequential damage occurred.

There were no incidents due to heart failure, and in a small number of cases it was necessary to re-hospitalize the patient for training.

The blood pressure values (systolic and diastolic) were measured by means of wrist gauges before, after half distance and at the end.

The systolic values decreased significantly by 11.6%. The resting blood pressure values measured at the beginning of each training session could confirm this trend: they also decreased by about 10%.

## Discussion

The results of the present pilot study show that training with a heart rate controlled pedelec leads to a measurable and significant benefit in patients with heart failure.

Cycling in general is one of the most suitable endurance sports for heart patients, since the body weight does not have to be carried.

However, it is used too rarely, especially by patients with heart failure. Especially for the group of people examined here, namely elderly people with heart disease, an individually dosed and moderate load according to medical guidelines is necessary. Electrically assisted cycling with a pedelec therefore represents a sensible and attractive alternative for these patients. However, it is still unknown whether and under which conditions a group of patients defined in this way will benefit from pedelec riding. It was also unclear whether this technology is safe enough to recommend the electric bicycle as a training device for cardiac patients in general and for those with heart failure. It should be noted that patients with heart failure who were previously deprived of physical activity are now recommended endurance sports with a high level of evidence and recommendation (IA), and that these sports provide a high benefit.

Studies such as the HF-Action study (7), Keteyian et al 2018 (8), the CROS study (9) conclude that the health benefit (mortality reduction) is considerable and is between 20 and 40%. For some of the parameters measured in this study (e.g. ejection fraction) this has also been proven in comparable studies. Erbs et al (10) report that the EF in a verum group of about 16 patients improved by 10.2% compared to a control group.

Studies on the special question of heart failure patients with the described experimental design are not yet available.

There are isolated indications (11) that the pedelec offers advantages compared to the “normal” bicycle and at least does not cause any loss of training.

The use of heart rate as a target and control variable is an adequate and recognized variable in endurance sports. The training effect can be read from it (12). This is because it shows itself in a higher load tolerance, an increasing peak VO2 and a decreasing heart rate at constant load. Adequate frequency control is therefore a desired prerequisite for such a training.

The present system HeartGo with an Android app on a commercially available smartphone meets this requirement in an optimal way. In the described training mode, the system allows the use of the heart rate as control variable. This works very well, even if the stability, especially that of the sensors, was in need of improvement. This can be clearly seen in the figures 1C and 1 D, which show that optimum frequency control is achieved over the entire training period.

The system was tested in a pilot project (MentorBike 4) (13) on patients in the sta-tionary rehabilitation phase II of a rehabilitation clinic and was judged to be practicable with high acceptance. However, the duration of the test was limited to three months, so it was appropriate to test and evaluate the app over a longer period of time.

The present study was designed as a 2-year pilot study with prospective design. Due to the low number of test persons, the results are only of limited significance. Nevertheless, studies with a low number of subjects lead to meaningful results, for example in the case of questions on physical activity in heart failure (19).

Parameters for objective recording of training data on the pedelec are not available at this time. In general, the training efficiency of heart groups has not been sufficiently investigated, although this physical training has been a central component of holistic rehabilitation sport for more than 40 years with a high level of acceptance. Buchwalski et al. (2002) (5) showed a considerable increase in performance by about 50%, but no effect on the “classic” risk factors. However, final statements about the validity of heart groups are missing. (14, 15). The safety of physical training in heart failure patients is classified as high, depending on the severity of the condition. However, it has been shown that the benefit clearly exceeds the risk (16). This could be confirmed in the study presented. In relation to the total number of 93 training units, the incident rate is 2.15%. In contrast, there is an average benefit of 28.6%, based on all significant changes (ergometry, ejection fraction, blood pressure behaviour, biomarkers).

For high-risk patients such as the group of test persons at the introduction of a pedelec concept, medical and paramedical supervision is necessary. The test persons should be thoroughly trained at the beginning of the training.

If we take as a measure of acceptance the sensation of exertion of the load up to 150 min or 50 km final load the sensation of load according to the BORG scale, this value remains constant at 11 until the end of the study. The ratio of distance (driving distance) to BORG value (18) then increases 2.5 times as an indication of a significant improvement in performance.

The clinical data such as increase in left ventricular ejection fraction LVEF, ergometry, 6-minute walking test (6MWD) (14,17), the decrease in the biomarker NT-pro-BNP or systolic blood pressure indicate a measurable improvement, even if a progression or therapy-related bias due to uncontrolled domestic activity or medically indicated changes in therapy cannot be excluded.

Further training effects of decrease in cardiovascular risk factors were discernible in the trend.

The decrease in heart rate and the increased pedaling load correspond to a moderate training effect (19), which also did not change substantially during the performance increase.

Some of the measured parameters, such as ejection fraction and the NTpro-BNP values, show an improvement in prognosis. However, hard endpoints require a longer observation period, as Taylor et al. 2019 (19) demonstrated.

Further studies with a larger number of test persons are therefore necessary for further clarification.

### Conclusion and Key messages

The described method of heart rate-controlled cycling on a pedelec in patients with heart disease is effective and acceptance was high. After careful instruction and practice, this form of training can also be recommended to patients for leisure time activities and training.

1. training of patients with moderate heart failure on the e-bike (pedelec) is possible, safe and improves the clinical condition of the existing underlying disease
2. a sustainable training effect for this group could be proven in the presented two-year pilot study.
3. the indicators of well-being and performance tolerance increased significantly and the blood pressure values decreased accordingly during the study period.
4. the objective performance parameters such as ergometry load and the 6-minute walking test improved.
5. the initially decreased left ventricular ejection fraction increased significantly.
6. the positive results of this pilot study with 10 subjects require verification by a multi-center follow-up study.

## Data Availability

The data availability is provided

https://herz-bike-saar.ghennersdorf.net/

